# Factors contributing to the emigration and retention of health workers in Bhutan

**DOI:** 10.1101/2025.10.05.25337373

**Authors:** Ugyen Tshering, Marjolein Dieleman

## Abstract

**Background:** Bhutan faces challenges maintaining an adequate health workforce, exacerbated by a post-COVID-19 exodus. Little is known about the reasons for emigration from Bhutan, known for its happiness and stable governance, and about mitigation strategies. This study aims to identify drivers of emigration and explore potential strategies for improving health worker retention.

**Method:** A narrative literature review was conducted in the context of Bhutan and similar settings, complemented by seven semi-structured interviews with researchers, policymakers, and health managers.

**Findings:** Bhutan has witnessed a surge in health worker departure post-COVID-19, particularly young professionals. Data from various sources underscore its gravity. Better financial opportunities, career prospects and a shift towards a materialistic mindset in Bhutanese community drive emigration. Societal pressure and family are both push and pull factors for health workers. Existing retention policies encompass financial incentives and bonds, but their impact remains to be evaluated. Drawing inspiration from global practices, strategies like circular/return migration, bilateral agreements, task shifting, and community engagement present promising avenues to mitigate emigration and fortify retention initiatives in Bhutan.

**Conclusion:** The study highlights the inevitability of emigration in a globalised world. While financial incentives and career opportunities are crucial, addressing societal factors and social values in retaining health workers is equally essential. By upholding the “right to move” while safeguarding the “right to health,” Bhutan can foster a sustainable and resilient health workforce.

## Background

Bhutan is a small landlocked country in South Asia, sharing its international borders with China in the north and India in the south (Fig 1)(1). With a total area of 38,394 km², Bhutan is one of the many small countries in the world, comparable in size to Switzerland. Bhutan’s projected population for 2022 was 756,129, with the fertility rate continued to remain below the replacement level of 2.1, which is a national concern. The proportion of people above 65 years is projected to increase from 6% in 2022 to 13% by 2047(2).

**Fig 1.**
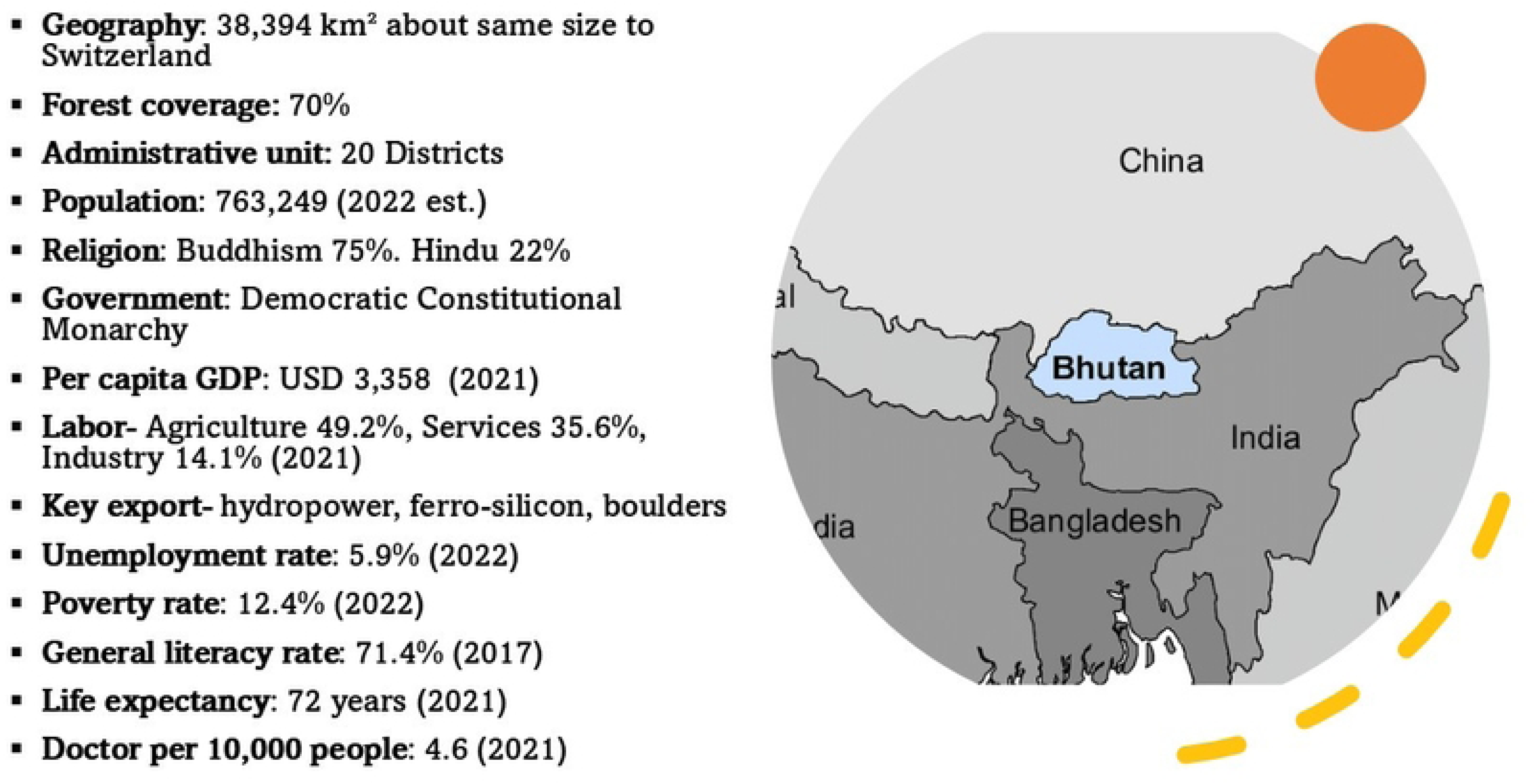
Bhutan in Brief. Adapted from National Statistical Bureau(131)

The health system is based on Primary Health Care (PHC) principles and is committed to achieving universal health coverage (UHC) by 2030. This pursuit of UHC is deeply embedded in the nation’s unique developmental philosophy of Gross National Happiness (GNH)(1,3,4). As enshrined in the Constitution, healthcare services are free to its citizens. Hence, the health system is predominantly financed and managed by the Government(5). The healthcare system is delivered through three-tier primary, secondary, and tertiary care(5–7).

In Bhutan, civil servants working in hospitals and health facilities constitute the human resources for health (HRH). The health workforce forms the second largest occupational group in the country’s civil service, comprising 16.3% of the total 30,194 civil servants in 2022(8).

Bhutan faces a significant challenge in ensuring an adequate supply of health workers to meet the healthcare needs of the population(5,6,9,10). This is attributed to the country’s limited capacity for health workforce production and inefficient human resource planning and deployment(10,11). In 2020–2021, the health workforce capacity was put to the test when the government brought back 50 doctors pursuing postgraduate training abroad, and engaged trainees from the medical university (KGUMSB) for COVID-19 management(12).

To compound the problem, retaining medical doctors and nurses in the health system has become a massive challenge for the country’s health system(13–15). Between 2011 and 2018, Bhutan lost 39 doctors, representing over 10% of the total doctors in the country(13). In 2022, a total of 2,646 civil servants departed from the civil service. Among them, health workers formed the second-largest group, with 290 individuals leaving, while an additional 74 took sabbatical/extraordinary leave (EOL)(8). A big concern is the number of doctors and nurses resigning or going on EOL.

This health workers emigration from Bhutan could be linked to several factors, which can be grouped into push and pull factors(16–19). Push factors refer to conditions that motivate individuals to leave a particular place or engagement, whereas pull factors encompass elements that attract individuals to a specific location or activity(20). Health workers might be leaving for better financial benefits, professional development, career advancement, and better quality of life(10,21,22). Understanding the interplay between different factors influencing the emigration decisions of health workers is crucial for effective policy formulation(23,24).

Addressing emigration more strategically in the country is critical to limit the strain placed on the health system already facing many challenges. Shortage and uneven distribution of health workers can lead to limited healthcare access, compromised quality of care, and poor performances, causing reduced productivity and increased burden on the existing health workforce(14,18,25–27). Ultimately, these factors contribute to poor health outcomes. Inadequate skills and numbers of health workers are attributed as one of the primary causes of patient safety issues in Bhutan(28,29). Moreover, health worker shortage results in increased inequities to healthcare access(5,11,30) and threatens public health, economic growth and development(18,31).

There are several published studies on the factors influencing migration and interventions to retain health workers in rural settings in low- and middle-income countries (LMICs)(26,32–35). However, there remains a gap in understanding why health workers emigrate from Bhutan(13–15), a country renowned for emphasising happiness and government stability(3,4). Therefore, this study aimed to identify and discuss factors contributing to emigration from Bhutan and explore potential strategies for improving health worker retention.

## Methods

We conducted a narrative literature review and complemented this with seven interviews.

### Narrative Literature Review

A comprehensive narrative literature review on HRH and health worker emigration was conducted in the context of Bhutan and similar settings (neighbouring countries from Nepal and India). Peer-reviewed articles were retrieved from PubMed, Embase, and Vrije University Library databases. In addition, search engines like Google and Google Scholar were also used to expand the literature search and include grey literature such as national reports, policy documents and publications from the MoH, WHO, World Bank, and other relevant organisations. A search strategy table showing the key words for the literature search is given in Annex 1.

The inclusion criteria focused on English peer-reviewed articles published from 2010 onwards, which corresponds to the adoption of the groundbreaking instrument, the Global Code of Practice on the International Recruitment of Health Personnel (WHO Global Code) by WHO member states(36,37) and the publication of the WHO guidelines on health workers attraction and retention in rural areas(38).

### Semi-structured Interviews with Key Informants

Seven SSIs with key informants (KIs) were conducted between May to July 2023. These interviews provided first-hand perspectives in obtaining a more nuanced understanding of the complex issues related to health worker migration and retention specifically for Bhutan. These included two policymakers, three health managers, and two academics.

### Data Collection and Quality Assurance

A topic guide included open-ended questions to explore the insights and experience based on KI’s professional capacities. It was pre-tested with a KI to assess their understanding of the questions. A transcript of this interview was discussed among researchers, and necessary changes were made. All interviews were conducted virtually through Zoom and recorded while both researcher and Kis spoke in English, one of the Bhutan’s official languages. Notes were also taken during the interviews. The duration of the interviews varied from 48-74 minutes. These interviews were transcribed verbatim by the researcher.

Both the researchers managed the entire data collection, processing, and analysis. The study received ethical clearance from the Research Ethics Committee, Royal Tropical Institute (KIT). Written informed consent was obtained from all KIs prior to the interview. Each KI was provided with an information sheet explaining the study objectives, procedures, risks and benefits, and given opportunity to ask questions. Consent forms were signed and dated by KI, and copies were retain securely by the research team. All information was derived, processed, and stored in encrypted folders on a password-protected computer. The identified themes and categories were shared with KIs, who gave critical feedback and suggestions to strengthen the analysis. Member checking was employed by sharing preliminary findings with participants to validate their contributions.

To ensure rigor and credibility, multiple quality assurance measures were applied. For the narrative literature review, sources were independently screened and selected according to predefined criteria, with disagreements resolved through discussion. For the interviews, apart from using the guide, and transcripts were cross-checked for accuracy. Data interpretation was conducted iteratively by the authors, with themes refined through consensus to enhance validity and minimize bias.

### Data Analysis

Deductive and inductive methods were applied to analyse both literature and interviews. A deductive approach was initially adopted to develop a coding framework based on the study objectives and pre-determined analytical framework. Three themes were identified: (i) Emigration trends and implications, (ii) Factors influencing emigration, and (iii) Retention strategies and policies. These themes served as the foundation for coding, encompassing seven factors as given in Table 1. NVivo 14 software was used to extract data from the interview transcripts using a coding system based on the identified themes and factors.

**Table 1.**
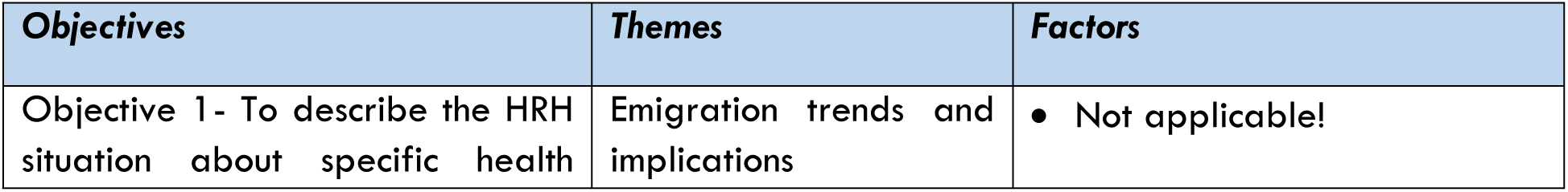
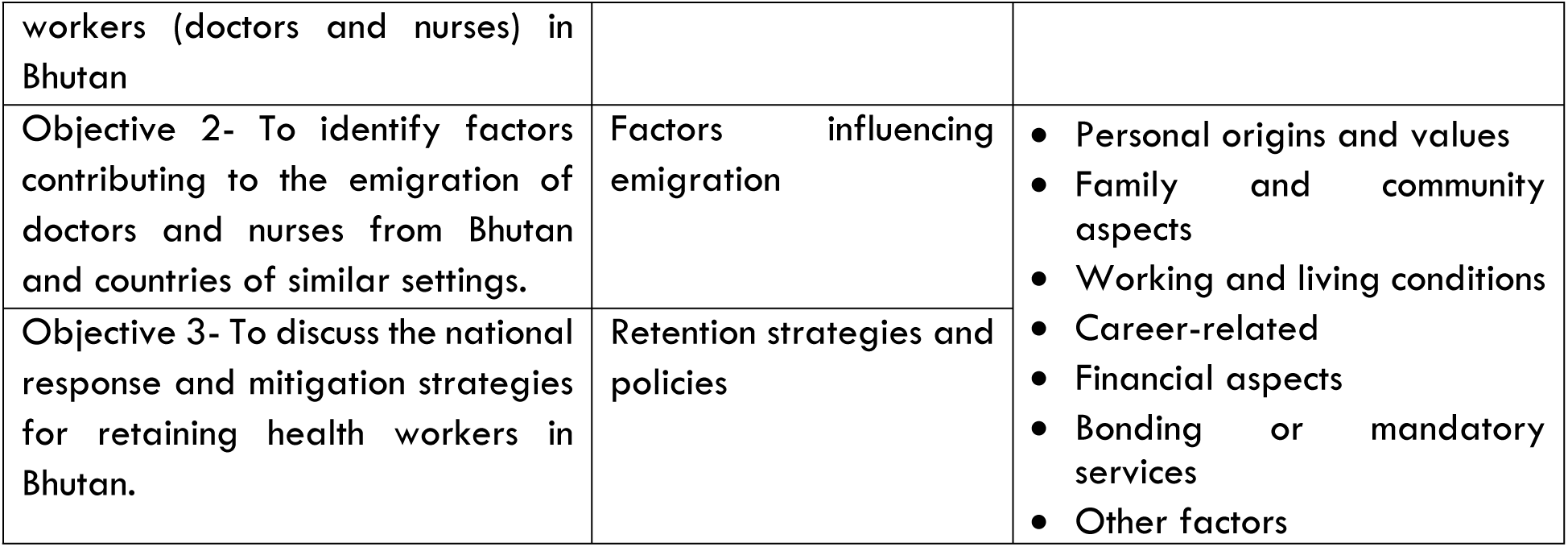
Classification of themes and factors (coding framework)

The interview transcripts and articles/ documents were systematically analysed by categorising them into themes and factors. During the process, the researcher identified and labelled new factors applicable to Bhutan’s context from the interviews.

### Analytical Framework

To guide the study, the framework from WHO’s guidelines on improving health workers retention in remote area(38) was used (Fig 2). This framework was originally adapted from Henderson and Tulloch’s framework on health worker’s motivation and retention(39).

**Fig 2.**
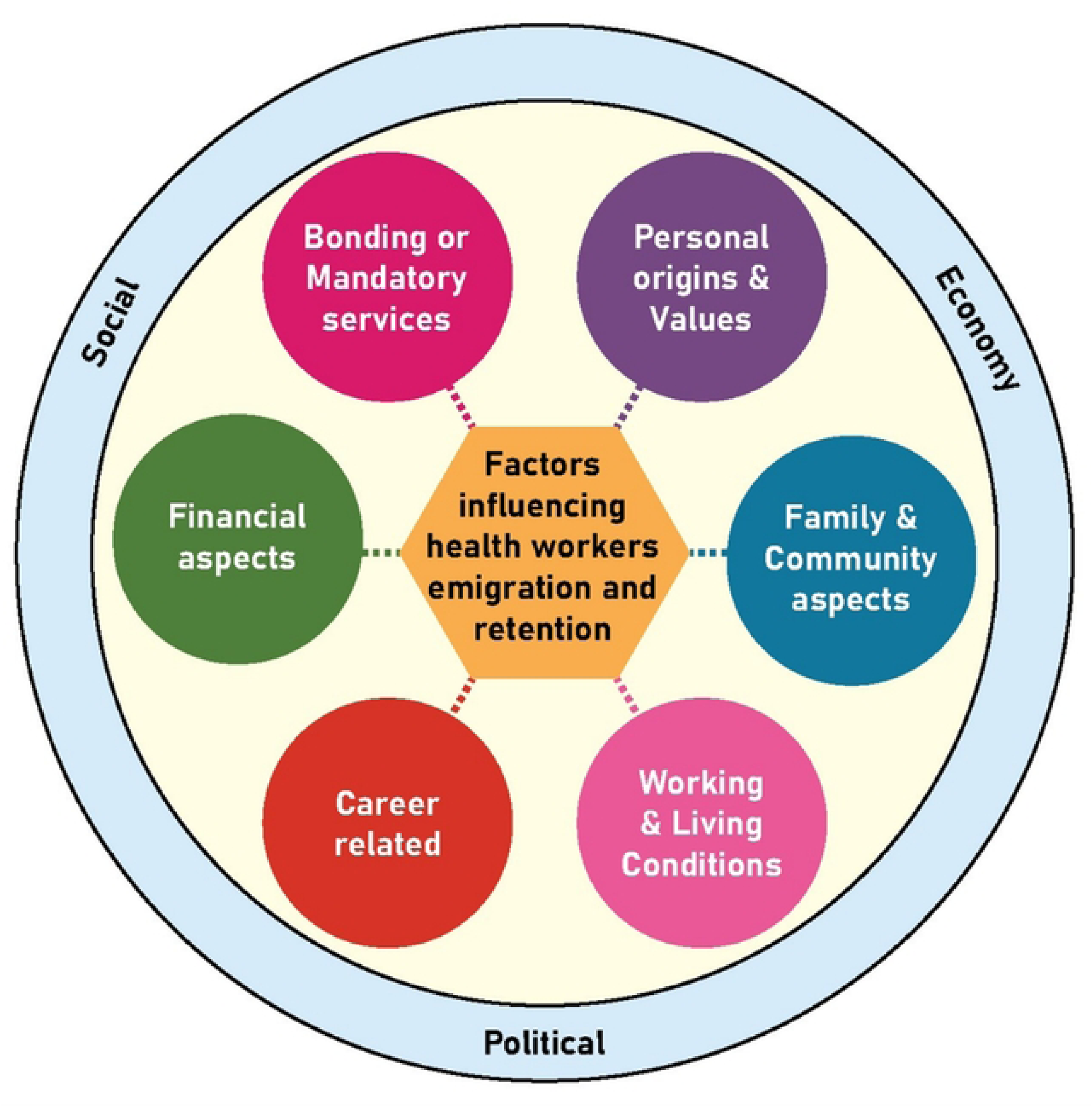
Analytical framework used for the study. Adapted from WHO guidelines 2010(38)

A slight modification of the framework was made prior to the data analysis. Initially representing the decision to stay or live rural area, the inner circle was replaced with “factors influencing health worker emigration and retention” to align with the study objective. The WHO framework has not addressed the broader social, economic, and political environment at the national and international levels, which significantly influences the migration and retention. However, this study acknowledges the need to consider the broader context to ensure a comprehensive understanding of the factors affecting migration and retention.

## Results

### HRH Situation, Trends of Emigration, and its implications

#### HRH status and stocks

In 2021, Bhutan had a total health workforce of 6643 including the support staff(40). This is tenfold increase from just 601 staff in 1985(5). There is now 1 health worker for approximately every 120 Bhutanese individuals. The doctor-to-population ratio stood at 4.62, and the nurse-to-population ratio was 20.9 (Table 2)(40), both falling below the required thresholds of 7.77 doctors and 58.64 nurses/midwives per 10,000 population for achieving the UHC Service coverage index(41). Similarly, the combined density of doctors, nurses and midwives (25.9/10,000 population) is also well below the indicative SDG threshold of 45/10,000 as set by the WHO (Fig 3)(30,42,43). In South-East Asia Region, the doctors’ density was lower than all countries except Indonesia while the nurses’ density was also lower than all other countries except Bangladesh(5). Nevertheless, nurses constitute the largest group of health workers in the country(44).

**Fig 3.**
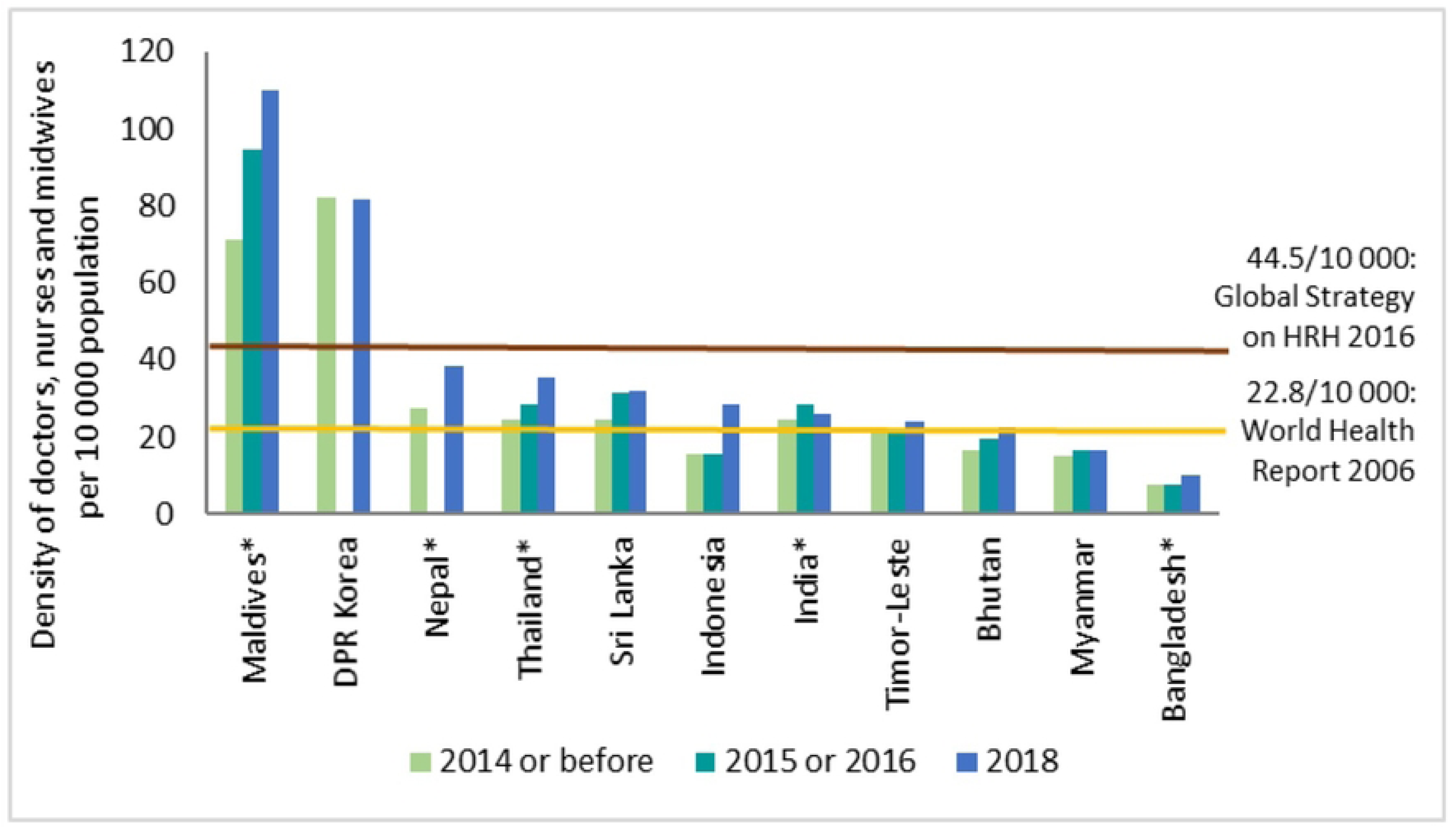
Trends showing the density of doctors, nurses, and midwives per 10,000 population in SEA Region countries, 2019. Source: WHO SEARO(45)

**Table 2.** Health workforce trend in last 5 years. Adapted from: Annual Health Bulletin 2022, MoH. (**40**)

| Sl no | Indicators | 2017 | 2018 | 2019 | 2020 | 2021 |
| --- | --- | --- | --- | --- | --- | --- |
| 1 | Number of Doctors* and density (per 10,000 population) | "345 (4.3)" | "337 (4.6)" | "318 (4.32)" | "336 (4.62)" | "354 (4.64)" |
| 2 | Number of Nurses** and density (per 10,000 population) | 1264 (16.2) | 1202 (16.5) | 1364 (18.6) | "1517 (20.9)" | "1608 (21.07)" |
| 3 | Number of Pharmacists and density (per 10,000 population) | "36 (0.5)" | "44 (0.6)" | "43 (0.6)" | "42 (0.6)" | "46 (0.6)" |
| 4 | Number of Health Assistants/ Clinical Officers, and density (per 10,000 population) | "636 (8.1)" | "604 (8.3)" | "620 (8.4)" | "650 (8.9)" | "683 (8.95)" |
| 5 | Number of Drungtshos (Indigenous physicians) and density (per 10,000 population) | "55 (0.7)" | "53 (0.7)" | "54 (0.7)" | "52 (0.7)" | "59 (0.77)" |
| 6 | Number of Sowa Menpas (Traditional medicine practitioners) and density (per 10,000 population) | "113 (1.4)" | "113 (1.5)" | "116 (1.6)" | "137 (1.9)" | "146 (1.9)" |
| 7 | Number and distribution of health facilities (per 10,000 population) | "276 (3.6)" | "279 (3.8)" | "288 (3.9)" | "289 (4.0)" | "289 (3.85)" |
| 8 | Ratio of nurses per Doctor (doctor to nurse ratio) | 3.6 | 3.5 | 4.3 | 4.5 | - |
| *includes both specialists and general doctors |  |  |  |  |  |  |
| **includes both clinical nurses and staff nurses |  |  |  |  |  |  |

Based on the Health Service & Human Resource Standard 2022-2026, projections estimate that Bhutan requires additional 195 doctors and 1,595 nurses to fill the gap by 2026(46,47). Looking at the workforce trend (Table 2) and annual supply projections (Annex 2), it will be a considerable challenge for the country to fill this daunting gap by 2026. Furthermore, in the upcoming year, the demand for doctors, nurses, and other health workers is expected to rise as Bhutan embarks on expanding new specialised health centres such as 500-bedded multi-disciplinary super-speciality hospital, and Royal Centre for Infectious Disease(47,48).

According to the interview, over the years, Bhutan has witnessed a notable increase in the diversity of health workers, including doctors, nutritionists, technologists, and nurses. All respondents recognised the ongoing critical situation in the health sector, driven by rising demands for healthcare services and concurrent health worker emigration. A manager expressed concern about the exodus of health workers can potentially undermine past achievements. Recent policy changes, like establishing cluster hospitals^i^ in certain districts, have led to specialists’ shortage, posing challenges for the larger regional referral hospitals, as mentioned by another manager.

> “The exodus of health workers in huge numbers from the system has the potential to undo so many things that we have achieved over the last six decades…”

> ∼{Respondent 1- Manager}

#### Recruitment and Training

Currently, the country doesn’t produce doctors. Bhutan still depends on neighbouring countries like India, Bangladesh, Thailand and Sri Lanka for medical education(33,49–51). Government selects top students from secondary schools and sends abroad to study medicine on various scholarship schemes. They are sent on the condition that they return to serve country after the graduation. Like other health workers, medical graduates must serve a duration two times the length of their course(5,50). Most return to the country to serve in civil service after graduation. As of December 2021, 471 students were studying health-related courses, including medicine abroad(52).

Nursing education had evolved since its inception in 1961, when Bhutan began developing its modern healthcare system. The Faculty of Nursing and Public Health under the KGUMSB played a significant role in nursing education(5,53). Establishing three private nursing colleges has increased the production capability of nurses in the country(54). Similarly, instituting the Faculty of Postgraduate Medicine under the KGUMSB in 2013 has contributed to addressing the shortage of specialists in Bhutan to a certain extent(49,53,55).

Every year, the graduates in the ‘Medical and Health Services’ category had the second highest intake into the civil service(8). The 2023 annual intake of doctors was only 27, while 126 nurses were recruited(56–58). The MoH also recruits health workers contractually in the middle of the year to cope with the demand(57). Despite the government’s concerted effort in health workers production, the intake of graduate-level health workers, particularly doctors, did not increase significantly. It is attributed to limited local health workforce production capacities and high dependency on the neighbouring countries for medical education(5,49,55). On a promising note, Bhutan just established its first MBBS programme at KGUMSB in 2023 (59), which is expected to enhance the country’s production capability of doctors.

#### Trends of Emigration

The exact size of the emigration remains unknown(10), as Bhutan’s health information management is disjointed and fragmented, preventing it from efficient data sharing and analysis(60,61). Estimations of emigration have varied widely, with different reports and media sources providing conflicting numbers. In this study, the data have been extracted from various sources.

According to the Department of Immigration, Bhutan has witnessed 50,125 migrants from Paro International Airport from January 2015 until March 2023 (Fig 4). This number is equivalent to one-seventh of the country’s population. In early 2023, the recorded number of emigrants was 5,000 per month(62,63). This figure does not include individuals who left through other land exits. From January 2018 and March 2023, 13,583 Bhutanese left for Australia via Paro Airport(62,63).

**Fig 4.**
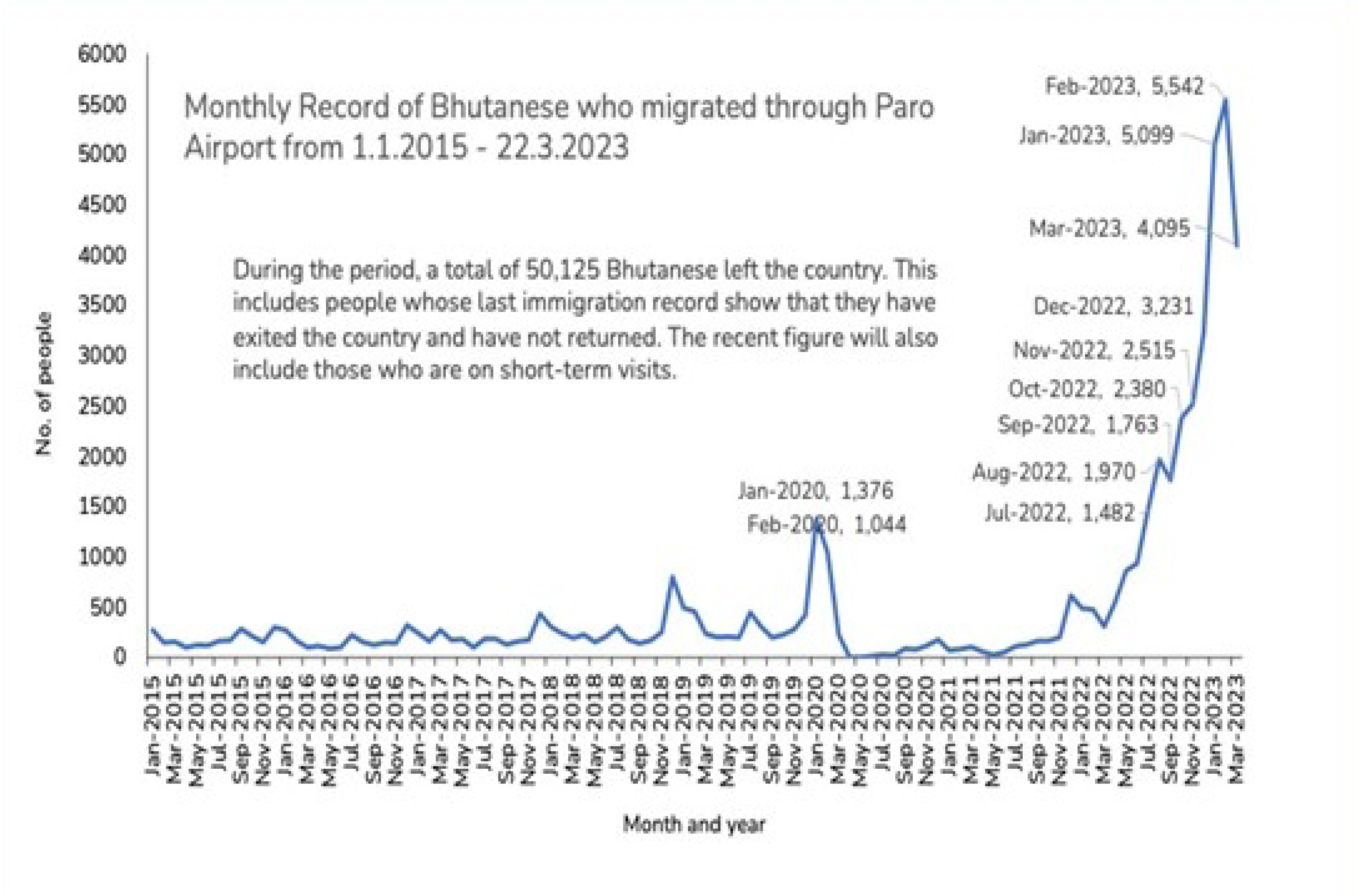
Monthly record of Bhutanese who left from Paro International Airport(63)

The median age of migrants to Australia was 28, while migrants to Kuwait was 24(62). This indicates that most migrants belong to the economically productive age group. This group comprises 5-10 years of working experience who play vital roles in developmental activities or service delivery(64). Regarding gender, females accounted for 51% of the total migrants(62).

As per the RCSC’s data series, there has been a noticeable shift in civil servants’ recruitment, resignations, and a net increase in 2022. There was a substantial increase in resignations (2,646), while the recruitment was 1791, resulting in a negative net increase of -855(8). This indicates a significant reduction in total number of civil servants and a potential challenge in retaining them.

The average number of resignations was 64 civil servants per month between January 2015 to May 2022. However, resignations rose to 234 per month from June 2022 onwards. A record-breaking 435 resignations was seen in January 2023(8,62). Further, 2,934 civil servants exited the system between January to June 2023, marking the highest attrition rate in recent years(65). The attrition rate in 2022 was 8.62%, but in the first six months of 2023, it surged to around 10%(8,65).

The health workers emigration from Bhutan has been a persistent phenomenon(13,66) and intensified notably after the COVID-19 pandemic(67). According to the MoH’s Attrition Report 2023, Bhutan has continued to witness an attrition rate of an average of 4% for health workers since 2018(58). The highest attrition rate (4.84%) occurred in 2022, with 223 health workers leaving the health sector (Fig 5), coinciding with the re-opening of international borders. Nurses have the highest attrition rate at 4.6%, followed by specialists at 2.5% and doctors at 1.7%(68). Nurses have consistently maintained the highest attrition rate since 2020 (Annex 3). In both the literature and interviews, it was evident that most health workers migrating from Bhutan are nurses.

**Fig 5.**
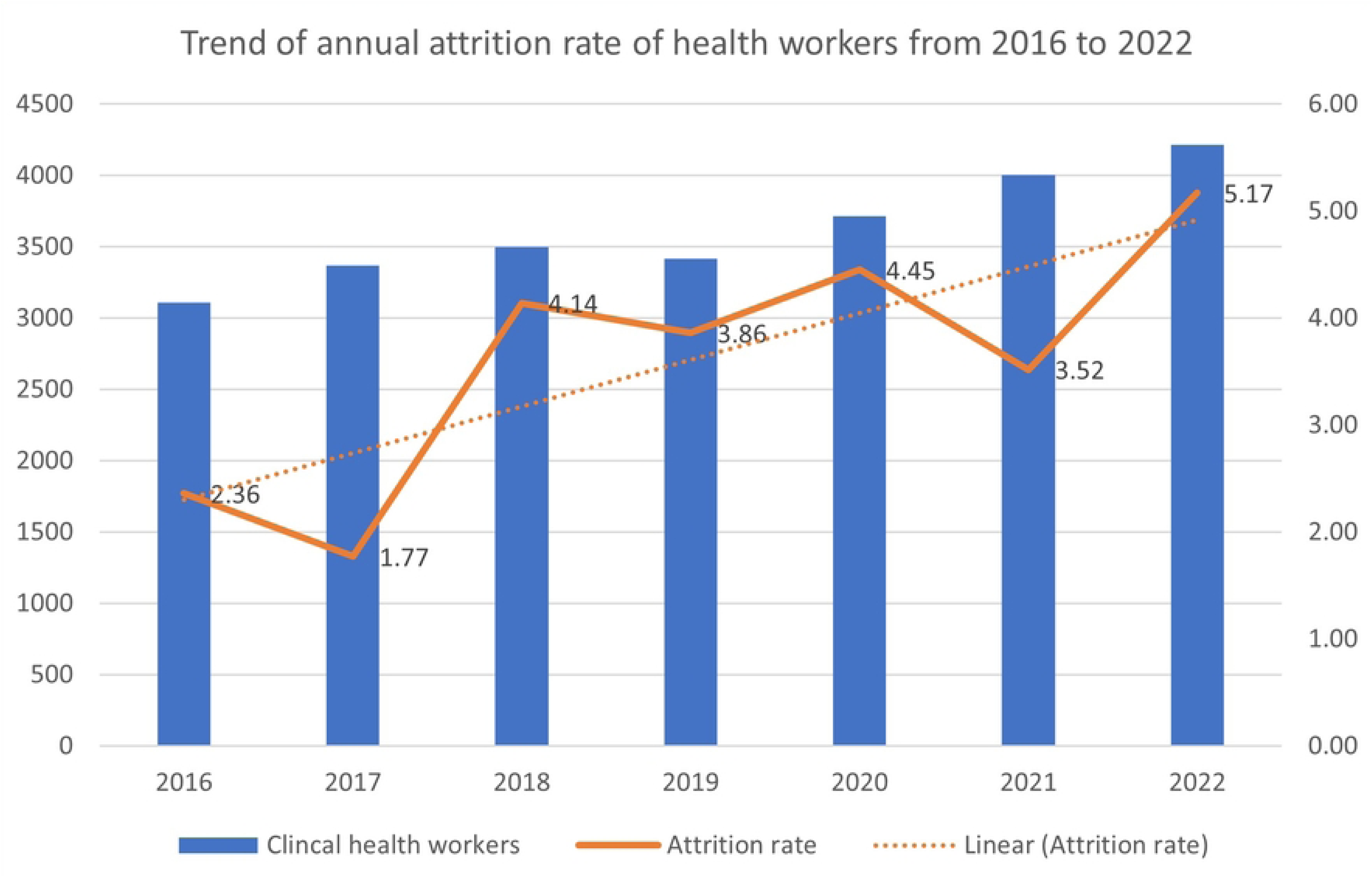
Trend of annual attrition rate for health workers from 2016 to 2022. Adapted from Attrition Report, MoH(58)

No official record exists for their whereabouts, but an ever-growing trend shows that many are migrating to Australia(62,69–71).

The interviews highlighted that some young doctors are increasingly preparing for opportunities abroad, particularly in countries like the United Kingdom (UK) and Australia. As highlighted by a manager, emigration predominantly occurs in early-career professionals with 7-8 years of working experience, posing a risk to meeting health workforce demands, especially in rural areas. Both literature and interviews consistently identify the United States (US), UK, Canada, and Australia as the top choices for health worker emigration(23,72–74).

> “… young, general physicians are not interested anymore to apply postgraduate within the country because most of them are busy preparing for opportunities in AMC Australia and the UK.”

> ∼{Respondent 1- Manager}

### Implications of Emigration

The exodus of health workers will have a huge impact at various levels, including the health sector, society, and family. In the health sector, it affects the quality of health service delivery(13,68). Moreover, the departure of skilled professionals poses challenges in finding immediate replacements, leading to burnout and depletion of workforce(64,75–77). At the societal level, the migration of youths and mid-level professionals will substantially impact on economic growth(64,71). And at the family level, both left-behind children and carers are more likely to suffer from mental disorders, depression and loneliness back home(78,79).

All respondents highlighted the similar implications of health worker migration. They added that the process burdens the remaining staff with increased responsibilities while remuneration remains unchanged, leading to dissatisfaction and prompting them to seek similar path. Additionally, respondents expressed concerns about a potential future crisis in competent leadership, which could jeopardise the functionality of hospitals and the overall healthcare system in Bhutan.

### Factors Contributing to the Emigration

#### i. Personal origins and values

In Bhutan, the traditional social values deeply rooted in Buddhist culture profoundly shape the lives of its people. Values such as *’ley jumdrey’* (cause and effect) and *’tha damtshig’* (sacred commitment) highlight the importance of interpersonal conduct, and social responsibility(80,81). Health workers were found to be motivated and satisfied by a moral duty to serve others, as indicated in studies(35,82). Such values can significantly affect their dedication and commitment towards the work and nation. In particular, there was a strong sense of patriotism in the past, focusing on serving the country and its people. However, in modern times, pursuing financial gain seems to be replacing these traditional values(66).

Academicians mentioned a shift towards materialistic mindsets in the Bhutanese community, which may influence the desire to emigrate while compromising spiritualism. Respondents also noted that modern societies are more vulnerable to external influences due to globalisation, decreasing individual resilience.

#### ii. Family and community aspects

Studies reported that family members play a crucial role in the decisions to migrate among Indian nurses. Nurses expressed the desire to migrate to better their families lives both at home and abroad(83). Additionally, family or relatives living overseas were an influential factor in health workers’ decision to migrate from India and Nepal(83,84).

Managers noted that health workers are drawn to countries like Australia for their children’s access to high-quality education and improved family prospect. They added that the prospect of gaining permanent residency adds to the appeal. Both policymakers and managers consistently reported the influence of peers and the domino effect. When health workers observe their colleagues leaving the country, achieving success, and improving livelihoods abroad, it triggers a peer-driven consideration of the same opportunity. This peer pressure creates a strong desire to seize similar prospects. Additionally, health workers were observed to emigrate while accompanying their spouses, a push factor contributing to the emigration.

> “Our parents are so committed to the point that they assure us not to worry about our children, as they will take care of them, and we should go to Australia when the opportunity is there.”

> ∼{Respondent 1- Manager}

#### iii. Working & living conditions

Poor working environments, such as a lack of essential medicines, equipment, and supplies for patient diagnosis and treatment, have influenced health workers’ decision to emigrate to LMICs(73,75,84). In Bhutan, lack of freedom in their roles was another significant contributing factor influencing the decision-making of health workers(69). The desire to leave is often attributed to heavy workload pressure and workplace stress in the case of Bhutan and Nepal(13,66,75,85). Health workers were found to experience stress due to the heavy workload caused by understaffing(86,87). Further, studies in both Bhutan and Nepal reported that the prospect of having better experience or exposure outside was one of the primary pull factors for health workers(69,88,89).

Similarly, respondents reflected that health workers face high work pressure, including prescriptive tasks and ad-hoc multitasking. This can negatively impact both their physical and mental health. An academician mentioned that delayed filling of vacancies in the health system burdens existing health workers with increased workload, causing further emigration.

The lack of a performance-based and proper feedback system influenced professionals to exit the system in Bhutan(69,88). The absence of such a system has also been identified as a demotivating factor for younger professionals(<35 years). Furthermore, they attributed emigration to dissatisfaction with office corruption or frustration with the system(69). In a 2019 study (N=147), only 64% of doctors were satisfied with their job in Bhutan(13). Another study (N=1009) revealed that doctors and nurses were reported to be the least satisfied professionals, with 62% and 63% satisfaction rates, respectively(15).

In the system, younger talents often struggle to come to light as seniors hinder their progress. There have been reports of nepotism, where individuals’ connections and family relationships play a significant role in career opportunities(66). Poor relationships with supervisors, lack of transparency, and job mismatch were also cited as reasons for emigration(66,88). A study from Bhutan emphasised the need for robust systems, including a strong work culture and monitoring system, to create a better future(14).

As part of the ongoing civil service reform, establishing of the National Medical Services (NMS) agency has separated clinical services management roles from the MoH(90,91). This move entrusts health workers, especially clinicians, with the stewardship of clinical areas, aiming to motivate and instil a greater sense of responsibility in carrying out their duties. Managers noted that the ongoing reform is crucial in bridging the gap between the central agency and field-level workers.

#### iv. Career-related

Lack of career mobility and advancement was identified as one of the main factors contributing to the health workers migration in South Asian countries(10,69,88,92). In Bhutan, limited upward mobility is a significant issue for professionals, given the scarcity of available executive positions(66). Moreover, since 2016, technical professionals like doctors, engineers and nurses were restricted from proceeding in the executive career pathway of the civil service until recently(14,93). This could have prevented them from continuing their careers in Bhutan.

In LMICs, a lack of learning opportunities, trainings and promotion was found to stimulate emigration decision(75). As per a study in Bhutan, 62% of the respondents cited the primary motivation for going to Australia as pursuing long-term training or studies(69). The absence of training and skill development opportunities was attributed to the emigration of Nepali nurses(86,89,92). Likewise, doctors expressed lower satisfaction with learning opportunities in Bhutan(13).

All respondents unanimously cited limited career growth and professional development as one of the main reasons for emigration. A manager noted that nurses’ career stagnation, which hinders growth and opportunities, prompts a desire to explore new paths.

> Nurses feel stagnant in their careers unless a divine intervention offers a chance for change.

> Therefore, they desire to explore different paths beyond their current profession…”

> ∼{Reference 4- Manager}

Studies highlighted the lack of appreciation and recognition contributing to heath workers’ poor performance(88,94,95). Many nurses expressed a perception of their work being undervalued(94). The interviews revealed that health workers feel undervalued and disrespected in Bhutan, leading to demotivation, and becoming significant push factors for emigration. This sentiment arises from a perceived lack of recognition and appreciation for their contributions to the healthcare sector.

The findings underscored the importance of good leadership and engagement in motivating Bhutanese health workers. They recognised that positive relationships with supervisors play a crucial role in retaining them within the system(88). The support and supervision from superiors, including the ministry, were found to be inadequate and poor(82). The need for effective bureaucratic leadership to instil a sense of dynamism within the organisation in Bhutan was equally emphasised(66). An academician emphasised that ineffective leadership, including poor communication and a lack of listening skills among leaders, hampers collaboration and a conducive work environment, contributing to push factors.

In March 2022, the RCSC managed out 47 executives who did not meet expectations during the leadership assessments(96). Several people emphasised that fear and job insecurity provoked civil servants to exit the system, as highlighted in national newspapers(66,97). Likewise, respondents acknowledged that the situation had instilled fear among the younger generation and may have repercussions on emigration trends.

#### v. Financial aspects

In many LMICs, poor remuneration and salary differentials have played a significant role in health workers emigration(18,21,23,75,92). Likewise, studies have identified inadequate salary and benefits as crucial factors for health worker’s emigration from Bhutan(66,69,88). In a notable statement, the former Prime Minister highlighted that public servants in Bhutan are paid significantly lower than in other countries, stressing the urgent need to address this disparity(97). Bhutanese professionals in Australia earned considerably more than their Bhutan-based counterparts, with some having monthly average incomes surpassing the annual earnings of entry-level civil servants in Bhutan(69). All respondents stated that the pull factors, particularly in countries like Australia, are strong due to the substantial difference in earning potential compared to working in Bhutan.

> “The pull factor is equally strong, not just the push factor. The salary difference is simply huge, and the earning potential abroad exceeds what one can earn in Bhutan in two to three years.”

> ∼{Respondent 1- Manager}

The study revealed that health workers were better paid, providing them with a compelling reason to stay(13). However, despite being the highest-paid civil servants in Bhutan, health workers continue to emigrate abroad(13,98,99). It supports that non-monetary incentives are equally important in improving motivation among professionals, as reported in the study(100). As we wrote this article, Bhutanese civil servants received another pay hike in July 2023(101).

#### vi. Bonding & mandatory services

Bonding is adopted in many LMICs to mitigate the migration of health workers(38,102,103). Bonding schemes offer a scholarship with a term-defined practice requirement upon completing studies(104). The duration varies from 1-9 years, depending on the country(102). Bhutanese health workers must serve for a period of two times the length of their courses(5,50). If doctors pursue specialisation or sub-specialisation, the obligation could become three or four times, respectively. Thus, a doctor is obligated for life-long to serve without the opportunity to explore different settings.

Despite lacking formal evaluation, most respondents find bonding programmes effective for staff retention. However, there are reservations regarding the strictness of the bonding and its potential impact on health workers’ commitment. An academician suggested that shortening postgraduate programmes from four to three years can both reduce bond and improve retention in Bhutan. According to a policymaker, if obligations with the RCSC did not bind doctors, there would be more consideration for leaving.

> “…Having that bond is also important because somehow they are obliged to serve the country since the government is spending on them. However, the obligation should not be enforced in an authoritarian manner.”

> ∼{Respondent 7- Policymaker}

#### vii. Other factors

In the context of health worker retention and emigration, it is evident that various significant factors extend beyond the ones identified in the analytical framework. These factors, including social, economic, and political aspects, will be explored in the subsequent sections.

##### a. Social factors

Bhutan’s unique societal values, which emphasise happiness/contentment over a materialistic world, may retain health workers to stay in the country(13,105). The support and encouragement from their family and spouses were the main reasons for motivation under social factor. Social respect and community support were additional motivating factors for their dedication to people’s welfare(82).

On the contrary, the situation seems to be changing. “Word-of-mouth”, coupled with societal pressure, played a significant role as early successful emigrants shared positive experiences, motivating more individuals to follow suit, resulting in a continuous increase in emigration(66). The interviews indicated that health workers face peer pressure and social expectations concerning emigration. Their families often compare their situation with friends and acquaintances who have migrated abroad. Respondents stressed that parents actively encourage their children to seek opportunities abroad, emphasizing the potential for higher income and a better future. The emigration trend, mainly to Australia, has become a common form of greeting, and staying back can make individuals feel undervalued or less successful, creating added pressure.

> “Frequent reminders and pressure from family members about friends doing well in Australia and earning significant amounts of money can influence the decision to explore opportunities abroad.”

> ∼{Respondent 1- Manager}

One intriguing finding from the respondents was the potential influence of forming de facto relationships (adopted spouses) contributing to emigration. De facto relationships pertain to partnerships where two individuals legally marry solely to travel to developed countries as dependents, despite not being in a genuine relationship. This phenomenon sheds light on another unique aspect of health worker emigration.

##### b. Political aspects

In many LMICs, political instability, economic collapse, and poverty have considerably influenced health workers’ decisions(75,106,107). Although Bhutan benefits from relative political stability with fewer extreme challenges, its health worker decisions are still affected by broader international political and economic conditions, and globalisation.

The demand for skilled health workers in HICs is a vital “pull factor” for emigration, offering better work conditions, higher salaries, and career advancement opportunities(21,66,75). The increase in demand for health workers in HICs is attributed to the ageing population, economic growth and failure to retain their medical graduates(42,74,104).

Globalisation is another crucial factor which exposes individuals to new opportunities and places Bhutan in competition with HICs for its employees(108). The migration trend is associated with the increased internet penetration and use, which exposes people to better opportunities abroad(66). Media portrayals depicting a high-quality life abroad have influenced emigration considerations in LMICs(21). Establishing the Australian and UK joint Visa application centre in Bhutan in 2016 has made visa applications more accessible, contributing to the emigration trend(66).

Likewise, the presence of several education consultancies in the capital city has led to applying for migration abroad, as stated by a manager. Recruitment agencies have a minimal role in recruiting health workers from the country, which is relieving for Bhutan. All respondents unanimously perceived that the favourable post-COVID-19 conditions, such as increased access to financial support, relaxed migration laws, and economic recovery, have fostered an enabling environment for considering emigration.

##### c. Economic situation

The overall economic situation of Bhutan plays a significant role in emigration, as it has deteriorated due to the COVID-19 pandemic and the effects of Ukraine’s war(109). At the same time, the unemployment rate has significantly risen in 2022 (5.9%) compared to 2017 (3.1%)(110), which could contribute factor to a spike in emigration(111). Along the same lines, policymakers and managers highlighted that the country’s economic challenges, particularly the aftermath of the COVID-19-induced economic downturn, have played a role. Bhutan’s current GDP per capita is around $3,000, which is considered low. Studies suggest emigration trends may reverse at $9,000 - $10,000 GDP per capita. Until then, professional emigration is likely to continue, added the manager.

### Retention Policies and Interventions

The RGoB has implemented various measures to address health worker retention, aiming to create a conducive environment that encourage them to stay and contribute to the country’s health system. However, the effectiveness of these initiatives has not been evaluated. These major interventions are described below in Table 3.

**Table 3.**
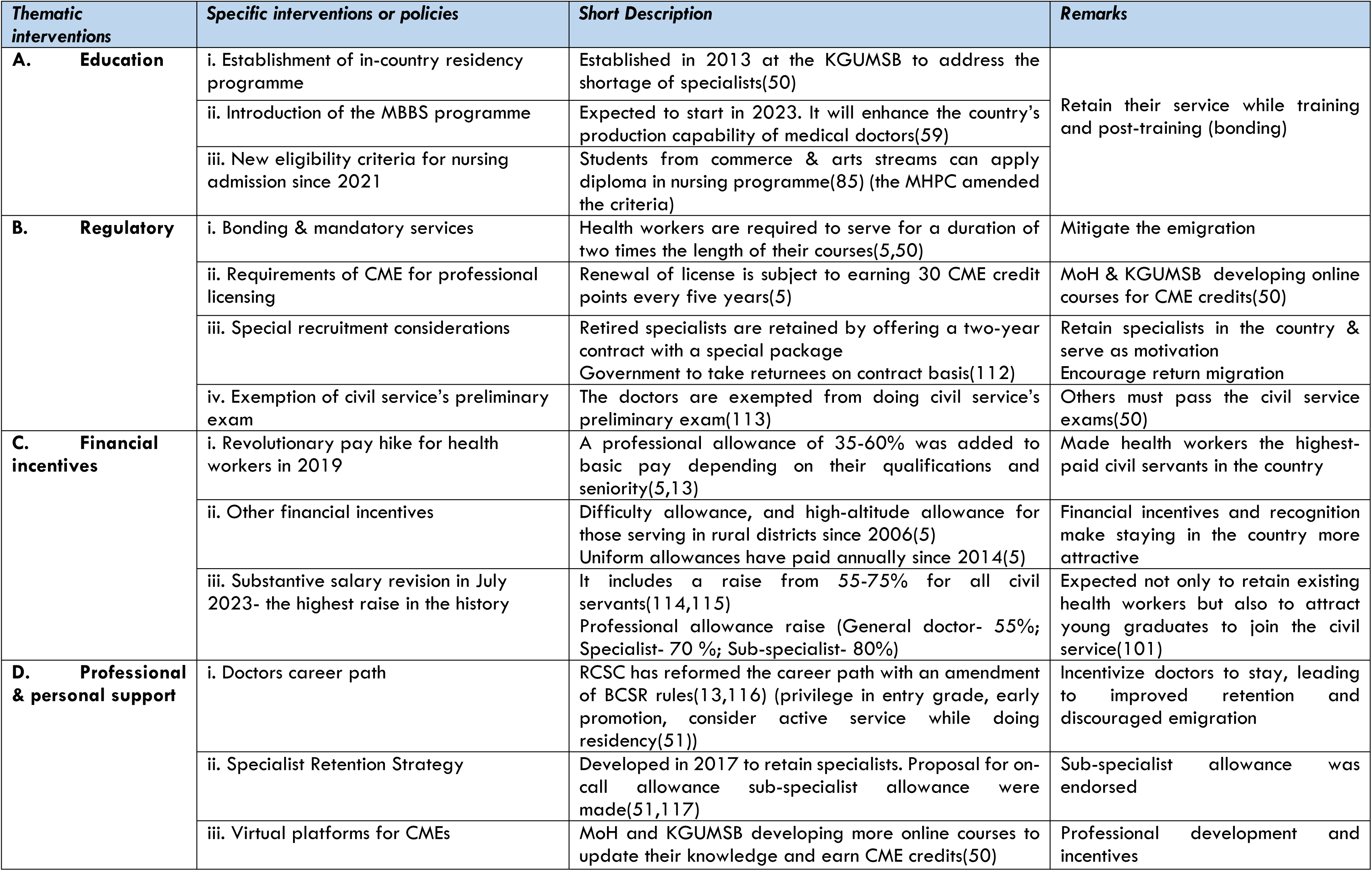
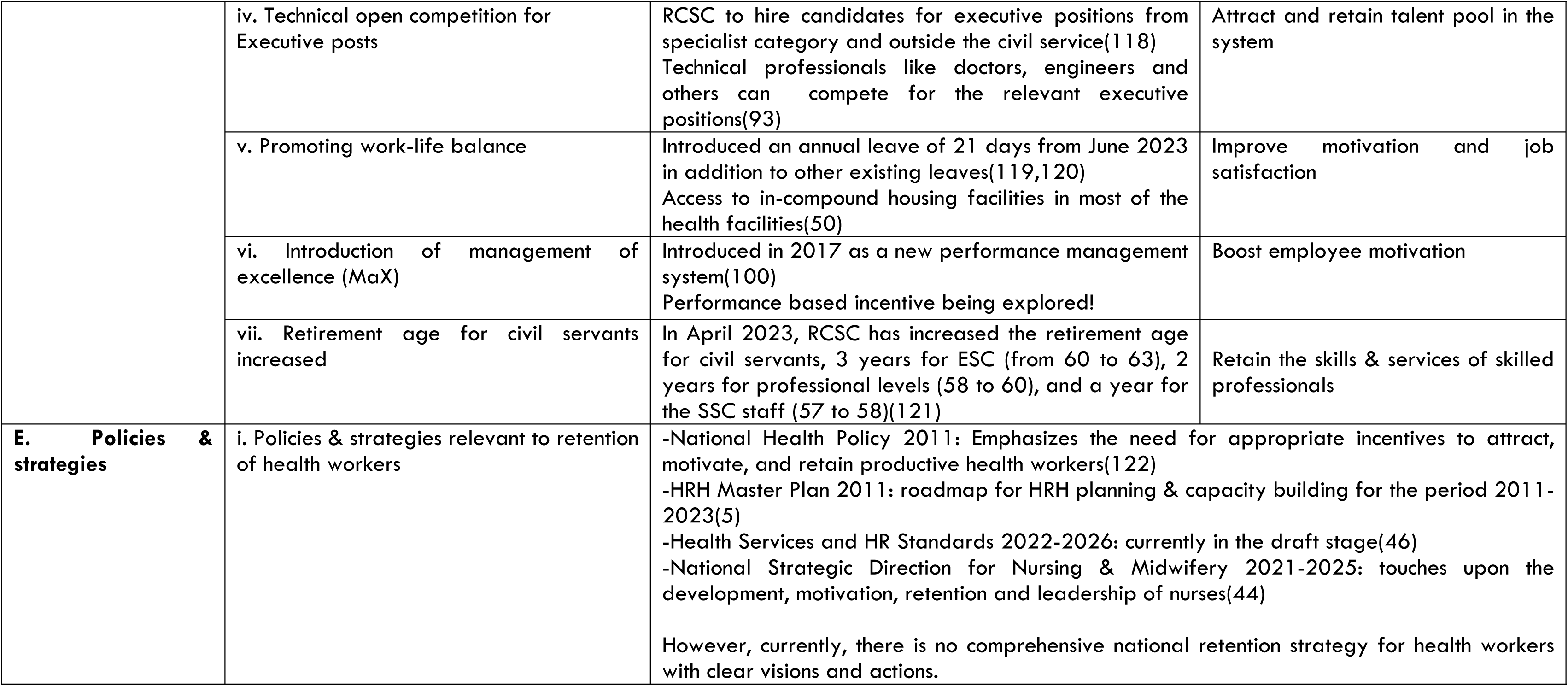
Interventions used to improve attraction and retention of health workers in Bhutan.

| **Thematic interventions** | **Specific interventions or policies** | **Short Description** | **Remarks** |
| --- | --- | --- | --- |
| **A. Education** | i. Establishment of in-country residency programme | Established in 2013 at the KGUMSB to address the shortage of specialists(50) | Retain their service while training and post-training (bonding) |
|  | ii. Introduction of the MBBS programme | Expected to start in 2023. It will enhance the country's production capability of medical doctors(59) |  |
|  | iii. New eligibility criteria for nursing admission since 2021 | Students from commerce & arts streams can apply diploma in nursing programme(85) (the MHPC amended the criteria) |  |
| **B. Regulatory** | i. Bonding & mandatory services | Health workers are required to serve for a duration of two times the length of their courses(5,50) | Mitigate the emigration |
|  | ii. Requirements of CME for professional licensing | Renewal of license is subject to earning 30 CME credit points every five years(5) | MoH & KGUMSB developing online courses for CME credits(50) |
|  | iii. Special recruitment considerations | Retired specialists are retained by offering a two-year contract with a special package<br>Government to take returnees on contract basis(112) | Retain specialists in the country & serve as motivation<br>Encourage return migration |
|  | iv. Exemption of civil service's preliminary exam | The doctors are exempted from doing civil service's preliminary exam(113) | Others must pass the civil service exams(50) |
| **C. Financial incentives** | i. Revolutionary pay hike for health workers in 2019 | A professional allowance of 35-60% was added to basic pay depending on their qualifications and seniority(5,13) | Made health workers the highest-paid civil servants in the country |
|  | ii. Other financial incentives | Difficulty allowance, and high-altitude allowance for those serving in rural districts since 2006(5)<br>Uniform allowances have paid annually since 2014(5) | Financial incentives and recognition make staying in the country more attractive |
|  | iii. Substantive salary revision in July 2023- the highest raise in the history | It includes a raise from 55-75% for all civil servants(114,115)<br>Professional allowance raise (General doctor- 55%; Specialist- 70 %; Sub-specialist- 80%) | Expected not only to retain existing health workers but also to attract young graduates to join the civil service(101) |
| **D. Professional & personal support** | i. Doctors career path | RCSC has reformed the career path with an amendment of BCSR rules(13,116) (privilege in entry grade, early promotion, consider active service while doing residency(51)) | Incentivize doctors to stay, leading to improved retention and discouraged emigration |
|  | ii. Specialist Retention Strategy | Developed in 2017 to retain specialists. Proposal for on-call allowance sub-specialist allowance were made(51,117) | Sub-specialist allowance was endorsed |
|  | iii. Virtual platforms for CMEs | MoH and KGUMSB developing more online courses to update their knowledge and earn CME credits(50) | Professional development and incentives |
|  | iv. Technical open competition for Executive posts | RCSC to hire candidates for executive positions from specialist category and outside the civil service(118)<br>Technical professionals like doctors, engineers and others can compete for the relevant executive positions(93) | Attract and retain talent pool in the system |
|  | v. Promoting work-life balance | Introduced an annual leave of 21 days from June 2023 in addition to other existing leaves(119,120)<br>Access to in-compound housing facilities in most of the health facilities(50) | Improve motivation and job satisfaction |
|  | vi. Introduction of management of excellence (MaX) | Introduced in 2017 as a new performance management system(100)<br>Performance based incentive being explored! | Boost employee motivation |
|  | vii. Retirement age for civil servants increased | In April 2023, RCSC has increased the retirement age for civil servants, 3 years for ESC (from 60 to 63), 2 years for professional levels (58 to 60), and a year for the SSC staff (57 to 58)(121) | Retain the skills & services of skilled professionals |
| **E. Policies & strategies** | i. Policies & strategies relevant to retention of health workers | -National Health Policy 2011: Emphasizes the need for appropriate incentives to attract, motivate, and retain productive health workers(122)<br>-HRH Master Plan 2011: roadmap for HRH planning & capacity building for the period 2011-2023(5)<br>-Health Services and HR Standards 2022-2026: currently in the draft stage(46)<br>-National Strategic Direction for Nursing & Midwifery 2021-2025: touches upon the development, motivation, retention and leadership of nurses(44)<br>However, currently, there is no comprehensive national retention strategy for health workers with clear visions and actions. |  |

## Discussion

### HRH Situation, Trends of Emigration and its Implications

The health workforce in Bhutan have evolved significantly over the years since the first introduction of modern healthcare in 1961. However, the current number of doctors and nurses still falls below the WHO’s recommended threshold of 4.45/1000 population. Bhutan continues to grapple with shortages of health workers, mainly due to maldistribution and the expansion of healthcare services. The significant emigration of doctors and nurses to HICs further exacerbates the situation, although precise figures remain unknown.

The presence of only one medical university in the country affects the production of an adequate number of medical graduates. Additionally, the high costs and the challenges of securing slots in foreign universities limit the intake for training. On the other hand, the local training capacities for nurses are considered adequate with the presence of public and private nursing colleges in the country.

The implications of emigration extend to multiple levels, including families, society, and the health sector. A noteworthy concern arises from the departure of the youths and mid-level professionals, which will likely impact the country’s overall economic growth considerably. However, if the current emigration trend continues, the country’s future prospects may appear less promising. It is crucial to recognise that this issue extends beyond mere comparisons of attrition rates or emigration numbers; it reflects a more profound challenge that requires careful consideration and thoughtful solutions.

The intersection of the “right to move” and the “right to health” significantly influences health worker retention and emigration dynamics. The “right to move” gives health workers the freedom to explore better opportunities and living conditions abroad, and it is unrealistic to expect emigration to come to a stop completely. At the same time, balancing this “right to move” with the “right to health” in Bhutan is crucial.

### Factors Contributing to Emigration

Various factors mentioned in the framework and literature are associated with motivation and satisfaction, which can encourage health workers to stay in the country or emigrate.

As evident from the findings, many health workers in Bhutan are driven to emigrate in pursuit of improved career prospects, higher remuneration, economic security, and improved living standards for their families. The allure of settling down and gaining permanent residency further adds to the appeal. These factors contribute to the brain drain phenomenon in Bhutan, similar to other LMICs.

Based on the findings, career-related aspects emerged as the key factors influencing health worker emigration. Lack of career advancement, inadequate learning opportunities, ineffective leadership, job security concerns, and poor recognition have driven them to seek better prospects abroad. This finding is consistent with previous studies conducted in other LMICs, where the absence of career progression has been linked to migration(76,123). To address this, policymakers should prioritise career growth opportunities, invest in CPD/CME programs, foster a culture of appreciation/recognition, strengthen leadership engagement, and create a conducive work environment. Implementing a performance-based and feedback system can also enhance job satisfaction and retention among health workers.

Among the various policy interventions, a significant focus was placed on improving the financial incentives for health workers as a strategic approach to retaining them. However, competing solely on financial incentives with developed countries presents challenges for Bhutan. Therefore, equal attention should be given to non-financial aspects, such as enhancing working conditions, promoting recognition and appreciation, mentorship opportunities, and performance-based incentives. Even in HIC like Ireland, retaining doctors proves challenging due to similar reasons(74,124). Hence, these complementary measures are essential in tandem with financial incentives to address the retention challenges effectively.

A concerning factor for Bhutan is the potential depletion of its intrinsic traditional values, such as social responsibility and contentment. There is a perceived shift towards more materialistic mindsets within the community, contributing to the desire to emigrate. For instance, despite health workers being the country’s highest-paid civil servants, with senior specialists earning more than the Cabinet Secretary(98), the highest position in the civil service, it is disheartening to witness such a trend. This reinforces the notion that financial incentives alone cannot be considered a panacea for the emigration issue.

The findings from the literature and interviews shed light on the influence of societal pressure, which is both a push and pull factor for health workers. On one hand, it may encourage individuals to pursue opportunities abroad for the promise of a better future, as illustrated in the “word-of-mouth” communication mentioned during the interviews. On the other hand, societal expectations might also pressure them to stay in the country despite personal aspirations for global exposure and professional growth. Similarly, the findings highlighted the significant influence of family members on migratory decisions, driven by the desire for stability in their families’ lives.

Social values, family dynamics, and societal influences are key factors impacting health worker retention in Bhutan, aligning with LMIC studies, particularly in African countries(20,21,25,34). However, current retention strategies primarily emphasize financial incentives and career prospects, overlooking these essential intangible aspects. This omission could result in insufficient emotional and personal engagement of health workers, a vital factor for a committed workforce and retention. To bridge this gap, Bhutan should actively promote social values, cultural appreciation, community engagement, and a sense of belongingness. Bhutan is set to introduce Gyalsung, the National Service training in 2024 mandatory for all youths attaining aged 18. This programme aims to cultivate sense of belongingness, instil social values, and foster life skills, potentially mitigating brain drain in the country(67).

The bonding requirement has been widely implemented in Bhutan. Despite the limited study on its effectiveness in the country, the study suggests that bonding programmes have successfully retained staff. This finding is consistent with evidence from LMICs and higher- and middle-income countries(38,102–104). However, there is a concern about the excessively stringent bonding scheme, potentially influencing health workers’ migration decisions.

On the other hand, the concept of “brain gain” is also worth considering in the context of HRH in Bhutan. “Brain gain” refers to the potential benefit of returnees to their home country with enhanced skills, ideas, and global exposure. As we address the challenges of “brain drain”, it becomes crucial to recognise the potential benefits of “brain gain” and develop strategies that encourage health workers to return to their homeland after gaining valuable experiences abroad through return or circular migration.

### Potential Mitigation Strategies

Based on the evidence and lessons gathered, it’s clear that a single intervention may not provide a sustainable solution for mitigating health worker emigration. Hence, a comprehensive approach is crucial, focusing on factors that retain health workers and evaluating policies that could hinder their return.

Return migration and circular migration present promising mitigation strategies. Return migration involves encouraging health workers who have gained international experience to return and contribute to their home country. By leveraging their expertise, skills, and cross-cultural exposure, they can positively impact the quality of healthcare delivery services. Initiatives like offering competitive salaries and conducive working conditions can encourage health workers to return to their homeland, fostering a mutually beneficial “brain gain” phenomenon.

However, returning health workers may experience difficulties in integrating with their community or profession, as reported in countries like India, Jamaica and Botswana(73,103,125). A study found that many employers and government representatives were unfamiliar with the concept of circular migration, leading to various barriers and challenges in its implementation(126). These obstacles include legal and administrative issues, lack of coordination, ethical concerns, skills recognition, etc. Therefore, identifying these barriers and exploring opportunities to encourage health workers’ homecoming is crucial. Conducive policies and well-designed support systems are essential in aiding and assisting the returnees in facilitating successful reintegration.

For instance, the existing BCSR pose a major obstacle, as health workers who have resigned are currently not eligible to rejoin. Besides, the age limit is 35 years to join the service. Many individuals may have gained valuable experience, but the government lack provisions to absorb them. However, the recent government announcement regarding the potential of allowing returnees to join civil service on contract gives us a promising ray of light(112).

Circular migration entails facilitating temporary work arrangements for health workers to gain international experience while maintaining strong ties with their home country. This can be achieved through bilateral agreements with other countries or participation in international health exchange programs. Drawing lessons from the Bhutan military’s successful engagement in the United Nations Peacekeeping mission(127), the health sector can implement similarly structured programs for health workers, offering international exposure and attractive incentives to encourage their return.

Bhutan can also draw insights from countries like Vietnam and Indonesia, where they have utilised bilateral G-to-G agreements with HICs, as mentioned earlier. These agreements can incorporate provisions for return migration, encouraging professionals to return home after overseas assignments. Moreover, adopting the WHO Global Code can streamline negotiations and advocacy efforts with HICs to establish such arrangements.

There is a growing discourse on privatising certain healthcare services in the country, owing to high demand and low growth in the public sector. This measure is seen as a means to reduce the burden on public hospitals, ensure quality healthcare services and retain health workers. A suggested approach is to permit health workers to partake in dual practice. Nonetheless, the evidence on the impact of dual practice on health workers retention is mixed and inconclusive(39,128,129).

Task shifting has emerged as a viable approach to tackle health workforce shortages. However, the current MHPC regulations hinder effective task shifting as they do not grant prescriptive rights to nurses. Despite this challenge, Bhutan can capitalize on existing Clinical Officers and Health Assistants as a strong foundation for addressing workforce shortages. These health workers’ cadres are less likely to emigrate as their qualifications are not fully recognised abroad. By strategically shifting tasks to existing Clinical Officers or introducing Nurse Practitioners, Bhutan can build on the system by empowering other professionals. This approach retains skilled workers and offers a sustainable solution for workforce challenges.

Lastly, the findings highlighted the importance of three broad factors: social, economic, and political aspects, that merit consideration in developing/adapting new analytical frameworks for understanding the subject matter. While the existing framework’s six factors offer valuable insights into individual motivations and local conditions, a comprehensive framework will encompass the broader systemic and global forces that influence health worker decisions. The decision to migrate or stay is not solely affected by personal factors or local circumstances but is interconnected with larger socio-economic and political contexts. For instance, better economic opportunities in source and destination countries can significantly impact health workers’ choices. Similarly, the global health labour market plays a pivotal role in shaping the emigration trend(107,130). By incorporating these broader aspects, the new framework can offer a more holistic understanding of health worker retention and emigration dynamics in Bhutan and similar settings.

### Strengths and Limitations

This study is a pioneering contribution to health worker emigration, being the first of its kind in the country. A mixed-method approach involving a thorough literature review and insightful KIIs offers a comprehensive and rich understanding of the subject. This study lays the groundwork for future research and serves as a reference for other researchers. The results hold crucial policy implications for health worker retention and emigration strategies. Furthermore, it contributes to the broader understanding of health worker migration in LMICs, informing similar studies in other settings.

Determining the precise extent of health worker emigration proved challenging due to the lack of official records and a fragmented health information system. Nevertheless, an estimate was gleaned from RCSC & MoH resignation records, offering a partial understanding of the emigration scale. The sampling methodology, utilizing KIs’ guidance and purposive sampling provides unique insights, but might have introduced bias. The selection process could overlook certain perspectives or individuals who could have provided valuable insights. Further, the small community dynamics may lead to participant homogeneity, restricting the representation of diverse perspectives and potentially overlooking the voices of underrepresented groups.

## Conclusions

The health workers emigration has become a longstanding and persistent phenomenon in our increasingly globalised world, and Bhutan is no exception. The post-COVID-19 period has witnessed a significant surge in the exodus of health workers, particularly young professionals, raising national concerns. While the exact magnitude of emigration remains unknown, the data from various sources underscore the gravity of this issue and its significance for the country. Evidently, this challenge extends beyond mere attrition rates or numerical comparisons; it calls for profound considerations and thoughtful solutions.

Acknowledging the “right to move” that allows health workers to seek better opportunities abroad is essential, and it is unrealistic to expect emigration to come to a stop completely. However, it is equally vital to balance this right with the “right to health” of the Bhutanese population. To strike this balance, Bhutan must adopt a comprehensive approach that addresses underlying factors contributing to emigration while implementing evidence-based strategies to improve health worker retention.

The study sheds light on the concerning trend of diminishing social values and sense of belongingness, and shifting materialistic mindset in the Bhutanese community. The study also highlights the influence of societal pressure and family members, which can be both a push and pull factor for health workers. Thus, giving equal emphasis on upholding social values while strengthening social support networks and family-related policies is crucial.

Since 2006, improved financial incentives have been a prioritized intervention to retain health workers, although its effectiveness remains unassessed. The existing gap emerges from the exclusive emphasis on financial incentives, overlooking crucial societal factors and social values. To address this gap, the government should adopt initiatives that foster community engagement, social values, and a sense of belongingness. Strengthening the emotional bond between health workers and their country can cultivate a committed healthcare workforce. Achieving this involves promoting cultural appreciation, recognizing contributions, and instilling pride among them.

To explore potential mitigation strategies, it becomes evident that a single intervention may not effectively address the complex issue of health worker emigration. Instead, a multi-sectorial and multifaceted approach tailored to the local context is crucial for developing sustainable retention strategies. Such an approach must not only consider financial incentives and career opportunities but also focus on addressing critical societal factors and social values among health workers.

Circular/return migration, G-to-G agreements, and task shifting are promising strategies to curb health worker emigration and enhance retention. Circular/return migration leverages the expertise of health workers with abroad experience, while G-to-G agreements safeguard their interests and facilitate international exposure. However, enforcing G-to-G agreements can be challenging due to their voluntary nature and lack of restrictions on recruitment agencies hiring for private companies. Task shifting is another critical approach for Bhutan’s health workforce challenges. By empowering and upskilling existing health workers, Bhutan can optimize its human resources and alleviate pressures leading to emigration.

In conclusion, tackling health worker emigration in Bhutan demands a comprehensive approach encompassing individual motivations, social influences, and broader socio-economic factors. This can be achieved by fostering social values, addressing societal factors, and offering non-financial incentives and career prospects. Embracing effective strategies like circular/return migration, G-to-G agreements, and task shifting will bolster retention, fortify the health system’s resilience, and ensure long-term sustainability.

## Data Availability

All relevant data are within the manuscript and its Supporting Information files.

## Acknowledgments

Our heartfelt gratitude to the colleagues at Royal Tropical Institute (KIT), The Netherlands for their unwavering guidance and support during the study. We are deeply grateful to all interviewees for their time and valuable insights, which enriched this research with firsthand experiences. Additionally, we would like to extend our appreciation to colleagues at MoH and the medical university (KGUMSB), Bhutan for providing access to essential data, which was crucial in shaping the study.

## Declarations

- Ethics approval and consent to participate- Attached
- Consent for publication- Not applicable
- Availability of data and materials- Not applicable
- Competing interests- Not applicable
- Funding- Not applicable
- Authors’ contributions

---

**Annex 1:** A search strategy table for literature search.

| **[Topic/object]**<br>**Problem/issue terms (OR)** |  | **[What I want to know about my topic/object?]**<br>**Factors-related terms (OR)** |  | **[Where is it?]**<br>**Geographical scope terms (OR)** |
| --- | --- | --- | --- | --- |
| “Human resources for health” | **AND** | Retention | **AND** | Asia |
| “Health workforce” |  | Migrat* |  | “South Asia” |
| HRH |  | Emigrat* |  | “South-East Asia” |
| “Health worker” |  | Attrition |  | Bhutan |
| “Healthcare worker” |  | “Brain drain” |  | “Developing count*” |
| “Health personal” |  | Motivation |  | LMICs |
| “Health professional” |  | Satisfaction |  | “Low- and middle-income count*” |
| “Medical doctor” |  | “Push and pull factors” |  |  |
| Doctor |  | “Barriers and enablers” |  |  |
| Physician |  | “Return migration” |  |  |
| Nurse |  | “Circular migration” |  |  |
|  |  | Performance |  |  |
|  |  | Resignation |  |  |
|  |  | Factors |  |  |
|  |  | “Health system” |  |  |
|  |  | “Working environment” |  |  |
|  |  | “Living conditions” |  |  |
|  |  | Social |  |  |
|  |  | Economy |  |  |
|  |  | Political |  |  |
| *: truncation symbol used to enable search |  |  |  |  |

**Annex 2:** Projection of annual supply of health workers. Source: Dept. of Adult & Higher Education and KGUMSB.

| **SI No** | **Course** | **2022** | **2023** | **2024** | **2025** | **2026** | **2027** | **Total** |
| --- | --- | --- | --- | --- | --- | --- | --- | --- |
| 1 | B. Entomology | 0 | 1 | 0 | 0 | 0 | 0 | 1 |
| 2 | B.Sc. Medical Lab Technology | 7 | 1 | 8 | 3 | 0 | 0 | 19 |
| 3 | B. Optometry | 0 | 0 | 2 | 0 | 0 | 0 | 2 |
| 4 | B. Pharmacy | 3 | 7 | 0 | 8 | 0 | 0 | 18 |
| 5 | B.Sc. Medical Imaging Technology | 2 | 0 | 3 | 1 | 0 | 0 | 6 |
| 6 | B.Sc. Physiotherapy | 0 | 5 | 2 | 2 | 0 | 0 | 9 |
| 7 | B.Sc Nutrition and Dietetics | 2 | 2 | 1 | 2 | 0 | 0 | 7 |
| 8 | B.Sc. OT Technology | 1 | 0 | 0 | 0 | 0 | 0 | 1 |
| 9 | B.Sc. Cardiovascular perfusion/CVT | 0 | 0 | 0 | 1 | 0 | 0 | 1 |
| 10 | **B.Sc. Nursing** | 28 | 36 | 46 | 79 | 0 | 0 | 189 |
| 11 | B.Sc. Dental Surgery | 0 | 0 | 0 | 2 | 0 | 0 | 2 |
| 12 | **MBBS** | 36 | 27 | 16 | 61 | 43 | 40 | 223 |
| 13 | Drungtsho | 8 | 7 | 8 | 8 |  |  | **31** |
| 14 | Menpa | 14 | 11 | 13 |  |  |  | **25** |
| 15 | **Staff Nurse (Diploma)** | 75 | 90 | 169 |  |  |  | **165** |
| 16 | Health Assistant | 20 | 25 | 25 |  |  |  | **45** |
| 17 | Dental Hygienist | 6 | 5 | 4 |  |  |  | **11** |
| 18 | Dental Technician | 6 | 5 | 5 |  |  |  | **11** |
| 19 | Laboratory Technician | 14 | 15 | 11 |  |  |  | 29 |
| 20 | Radiographer | 10 | 10 | 10 |  |  |  | 20 |
| 21 | Pharmacy Technician | 10 | 16 | 8 |  |  |  | 26 |
| 22 | Physiotherapy Technician | 5 | 5 | 8 |  |  |  | 10 |
| 23 | EMR | 21 | 20 | 21 |  |  |  | 41 |
| 24 | ENT Technician | 2 | 6 | 1 |  |  |  | 8 |
| 25 | OT Technician | 4 | 5 | 1 |  |  |  | 9 |
| 26 | Ortho Technician | 3 | 5 | 2 |  |  |  | 8 |
| 27 | Ophthalmic Technician | 4 | 8 | 5 |  |  |  | 12 |
|  | **Total** | **281** | **312** | **86** | **167** | **43** | **40** | **929** |

**Annex 3:** Trend of attrition rate of health workers in selected categories from 2020 to May 2023. Adapted from the Annual Attrition Report, MoH.

| Year | Staff Category | Total Employees | Total Recruitment | No. of separation | Average employees per Year | Attrition Rate (No. of separations /average employee*100) |
| --- | --- | --- | --- | --- | --- | --- |
| 2020 | Doctor (incl. Specialist) | 336 | 22 | 3 | 346 | 0.87 |
|  | Dental Surgeon and Specialist | 70 | 6 | 1 | 73 | 1.37 |
|  | Nurse | 1549 | 227 | 68 | 1629 | 4.17 |
|  | Laboratory Officer and Technician | 339 | 22 | 3 | 349 | 0.86 |
|  | Pharmacist and Pharmacy Technician | 226 | 32 | 12 | 236 | 5.08 |
|  | Health Assistant | 650 | 52 | 31 | 661 | 4.69 |
| 2021 | Doctor (incl. Specialist) | 379 | 47 | 14 | 396 | 3.54 |
|  | Dental Surgeon and Specialist | 73 | 5 | 2 | 75 | 2.67 |
|  | Nurse | 1 634 | 177 | 81 | 1 682 | 4.82 |
|  | Laboratory Officer and Technician | 348 | 33 | 10 | 360 | 2.78 |
|  | Pharmacist and Pharmacy Technician | 237 | 18 | 4 | 244 | 1.64 |
|  | Health Assistant | 655 | 28 | 2 | 668 | 0.30 |
| 2022 | Doctor (incl. Specialist) | 396 | 36 | 14 | 407 | 3.44 |
|  | Dental Surgeon and Specialist | 76 | 2 | 4 | 75 | 5.33 |
|  | Nurse | 1 644 | 72 | 66 | 1 647 | 4.01 |
|  | Laboratory Officer and Technician | 361 | 10 | 14 | 359 | 3.90 |
|  | Pharmacist and Pharmacy Technician | 251 | 15 | 3 | 257 | 1.17 |
|  | Health Assistant | 661 | 31 | 14 | 669 | 2.09 |
| 2023 | Doctor (incl. Specialist) | 388 | 35 | 7 | 402 | 1.74 |
|  | Dental Surgeon and Specialist | 77 | 6 | 2 | 79 | 2.53 |
|  | Nurse | 1 657 | 131 | 79 | 1 683 | 4.69 |
|  | Laboratory Officer and Technician | 366 | 20 | 15 | 369 | 4.07 |
|  | Pharmacist and Pharmacy Technician | 253 | 12 | 4 | 257 | 1.56 |
|  | Health Assistant | 667 | 27 | 14 | 674 | 2.08 |

## Footnotes

^i^central hospital equipped with 5 specialists(gynaecologist, paediatrician, general surgeon, anaesthesiologist, and medical specialist) and caters to two or more districts.

